# Behavioural activation for low mood and anxiety in male frontline NHS workers (BALM): a pre-post intervention study

**DOI:** 10.1101/2025.04.17.25325391

**Authors:** Paul Galdas, Della Bailey, Steve Bell, Kate Bosanquet, Carolyn Chew Graham, David Ekers, Simon Gilbody, Liz Littlewood, Michael Mawhinney, Heidi Stevens, Katie Webb, Dean McMillan

## Abstract

**Objectives:** To evaluate the impact and acceptability of a tailored, gender-responsive behavioural activation intervention for improving depression and anxiety in male NHS frontline workers.

**Design:** Pre-post intervention study.

**Setting:** Three National Health Service (NHS) organisations in the North of England.

**Participants:** Forty-five men aged *≥*18 years working in a frontline NHS role scoring in the subclinical range (5-14) on the PHQ-9 (depression) and/or the GAD-7 (anxiety) at baseline.

**Interventions:** A tailored Behavioural Activation treatment programme consisting of up to eight telephone support sessions over a period of 4–6 weeks, accompanied by a Behavioural Activation self-help manual.

**Main outcome measures:** Self-reported symptom severity of depression, assessed by PHQ-9, and anxiety, assessed by GAD-7, at baseline and 4- and 6-months. Acceptability from the perspectives of male study participants and coaches who delivered the intervention was assessed in a nested qualitative study using the theoretical framework of acceptability (TFA).

**Results:** PHQ-9 and GAD-7 scores decreased from baseline to 4 months on both the PHQ-9 and GAD-7. While scores increased from 4 months to 6 months, the 6-month scores remained below those of the baseline scores. Acceptability of the intervention was high across all constructs of the TFA. The practical and action-oriented strategies of the intervention, and the confidential, flexible, convenient mode of delivery, worked to support men’s engagement with the intervention.

**Conclusions:** Delivery of a tailored, gender-responsive Behavioural Activation intervention was appealing to, and beneficial for, men working in frontline NHS roles with less severe depression and anxiety. The BALM intervention offers promise as a tailored workplace mental health programme that is aligned with men’s needs and preferences and can help overcome a reticence to engage with mental health support in NHS staff and beyond.

**Trial registration:** ISRCTN 48636092

**Strengths and limitations of this study:**

- This is the first study to evaluate the impact and acceptability of a Behavioural Activation intervention for improving depression and anxiety in men using a gendered lens, contributing novel findings of relevance wider than the study context.
- The use of the TFA to inform data collection and analysis enabled a theoretically informed, systematic evaluation of the acceptability of the intervention.
- Rich data were gathered from qualitative interviews with a large sample of male participants and coaches who supported intervention delivery, including participant non-completers.
- Participants were a self-selecting sample of men who may have been particularly motivated to take part in the intervention. It is possible their views on the acceptability of the Behavioural Activation (BALM) intervention are more positive than the wider population of male NHS staff. Our sample also lacked ethnocultural diversity and representation of some NHS staff groups, particularly ancillary roles, that limits the transferability of the findings.
- The quasi-experimental research design is not sufficiently reliable to make causal inferences.

## Introduction

There are unprecedented and sustained high levels of sickness absence in the National Health Service (NHS) workforce in England, with increases observed across all staff groups since 2009. Rates rose sharply during the coronavirus disease 2019 (COVID-19) pandemic and have remained elevated compared to pre-pandemic levels.^1^

The major cause of sickness absence in NHS staff are mental health conditions, particularly anxiety and depression, which now account for around a quarter of all sick days.^1^ ^2^ Approximately 6 million sick days for mental health and wellbeing related reasons were reported in 2022, an increase of 26% since 2019.^2^ By contrast, mental illness accounts for 8%–12% of sickness absence each year in the broader UK labour force, meaning NHS employees are two to three times more likely to require sickness absence leave for mental ill health than the average UK employee.^1^ Sickness absence of health care professionals can lead to suboptimal patient care and significant increased costs, and is increasingly a cause for staff to leave the NHS.^1^ ^3^ As a result, there is growing interest in interventions to support the mental health and wellbeing of health care workers, globally.^3^

Frontline health care staff are exposed to a variety of workplace stressors throughout their careers that can lead to mental distress.^4^ Causes vary, but high workload, long hours, difficult shift patterns, and the absence of healthy rotation schedules that accommodate adequate rest, sleep, and restoration over time have been consistently identified in the empirical literature as important contributors to much of the common mental health challenges faced by health care professionals.^4^ ^5^ This distress and these adverse psychological outcomes do not typically require high-intensity mental health interventions.^6^ A number of ‘low-intensity’ strategies have been recommended; these include supporting regular breaks, adequate rest and sleep, a healthy diet, physical activity, peer and family support, avoidance of unhelpful coping strategies (e.g. use of alcohol and drugs), limitation of social media use, mindfulness and meditation practices, and professional counselling or psychological services.^3^ ^5^ However, uncertainty exists about the best models of care to effect positive change in this population. Thus far, workplace interventions have failed to produce a continuous downward trend in sickness absence rates.^7^ Barriers to uptake and engagement have also been reported; in particular, frontline workers, or the organisations in which they work, not being fully aware of what they need to support their mental well-being.^5^

Relevant here is the culture in many frontline health care and emergency professions that has promoted the idea that they are (or should be) stronger and more resilient than the general population.^8^ This ethos can impede the willingness of staff to acknowledge psychological problems and seek support, and may be particularly problematic for male frontline workers by adding to the stigma of help seeking for mental health concerns found in the general population.^8^ ^9^ Traditional masculine norms characterized by self-reliance, stoicism, and restrictive emotionality are associated with higher stigmatization of mental health problems and reluctance to acknowledge or seek help from professional mental health services.^10^ Although the tailoring of psychological support to address these issues has been a long-standing topic of concern in the men’s health literature, little evidence exists on effective gender-responsive interventions (i.e. those which pay attention to men’s specific needs, gender-based barriers, and gender differences) in the general workforce or frontline health care communities.^8^ ^11^ To address this gap in the evidence, the current study aimed to develop, deliver, and evaluate a gender-responsive mental health intervention for low mood and anxiety in male frontline NHS workers, based on Behavioural Activation (BA) principles.

BA is a brief, evidence-based, guided self-help treatment that offers promise as a gender-responsive approach to early mental health intervention for men due to its practical, collaborative, and action-oriented strategies that are consistent with a strengths-based masculinities approach that reinforces participants’ sense of autonomy, control and independence.^11^ ^12^ The evidence for use of BA as an early intervention in people without a formal mental health diagnosis is strong. National Institute for Health and Care Excellence (NICE) guidance in the UK^13^ recommends BA as a first line treatment option for adults with less severe depressive symptoms (i.e. mild depression and those with ‘subthreshold’ symptoms which fall below the criteria for diagnosis) and significant effects have also been reported for improving quality of life and anxiety.^14^ However, to our knowledge, no study has evaluated the impact and acceptability of BA as a workplace mental health intervention for men in either the general population or in frontline health care staff.

The specific objectives of this study were to: (1) evaluate the impact of a tailored, gender-responsive BA intervention for improving depression and anxiety in male NHS frontline workers; and (2) explore the acceptability of the intervention from the perspective of male study participants and the coaches who delivered it.

## Methods

### Study design

We used a single sample, pre–post study design to assess baseline depression and anxiety measures in male frontline NHS workers and determine the impact of the intervention on their mental health at 4- and 6-months, with a nested qualitative study using semi-structured interviews to explore the acceptability of the intervention from the perspectives of male study participants and the coaches who delivered it.

### Participants and sample

Participants were recruited from three NHS organisations (‘Trusts’) in the North of England: two hospital Trusts and an ambulance service. To be eligible for the study, participants had to identify as male, be aged *≥*18 years, and work in a ‘frontline’ NHS role as defined in NHS Workforce Statistics.^15^ Eligible staff groups included professionally qualified clinical staff, support to clinical staff, and NHS infrastructure support staff.

Participation was voluntary and selected as a convenience cluster sample. Study information was distributed via posters displayed in staff areas and electronic bulletins/staff intranet across the three study sites, and through social media platforms. Potential participants were invited to visit a secure online site to complete a consent form if they were interested in taking part after reading study materials, followed by a baseline questionnaire assessing their eligibility. As BALM was an early mental health intervention, individuals were eligible for participation in this study if they scored in the subclinical range (5-14) on the Patient Health Questionnaire-9 (PHQ-9) and/or the Generalised Anxiety Disorder-7 questionnaire (GAD-7) at baseline. Individuals scoring outside this range, currently receiving treatment for a mental health condition, or who had been previously diagnosed with bipolar disorder, schizophrenia, or other psychotic disorder, were excluded.

Following completion of the baseline questionnaire, eligible participants were sent a letter via email to confirm their participation. Those who did not meet the eligibility criteria were sent a letter thanking them for their interest in the study, including signposting to other mental health and wellbeing support programmes, or referred to their GP, as appropriate.

### Intervention

The intervention, ‘BALM’, was a structured programme of Behavioural Activation tailored to specifically meet the needs of male NHS workers with less severe depression and/or anxiety symptoms, through the application of a gendered lens.^16^ BA is a practical treatment that explores how life events can result in a reduction of meaningful activity, which in turn leads to feelings of low mood and anxiety. In response to these feelings, a person may attempt to cope through avoidance. While this may work in the short-term, in the long-term it may lead to further reductions in important or meaningful activity. In the current study, life events were principally related to both workplace stressors, such as high workload, long hours, and difficult shift patterns, but could also include non-workplace stressors such as changes in routine or life circumstances, relationships, and financial pressures. The intervention consisted of a coach and participant working together to develop a collaborative treatment plan that sought to increase meaningful activities and reduce avoidance.

The process of intervention tailoring for BALM involved a stakeholder advisory group comprising NHS psychological wellbeing practitioners and male NHS workers from medicine, paramedicine, nursing, allied health, and ancillary occupational groups. The group met on three occasions in co-design workshops to tailor aspects of the intervention relating to context, content, and communication in accordance with concepts outlined in the 5C Framework for designing men’s health programmes.^16^ The main adaptations related to the format, design, content, and use of language in the self-help manual, and the content of the coach training to incorporate evidence-based guidance on ways to engage men in mental health support.

The final BALM intervention consisted of up to eight telephone or video call support sessions (according to participant preference) over a period of 4–6 weeks, accompanied by a BALM Behavioural Activation self-help manual. Sessions lasted approximately 30 minutes and were facilitated by coaches voluntarily recruited from the three NHS Trust study sites. Both male and female coaches were recruited from a broad range of clinical and non-clinical NHS backgrounds including psychological well-being practitioners, nurses, paramedics, research assistants, clinical support officers, and crisis support workers. Coaches were trained to facilitate the intervention and review progress and outcomes in three online full-day workshops. In response to feedback in the co-design workshops, BALM participants were allocated a coach from a different NHS employer to their own to help to overcome barriers to engagement by maintaining confidentiality and anonymity. A secure computer system was used to monitor care, and supervision of coaches was provided by D.M. and D.B.

### Outcomes

We studied outcomes of self-reported symptom severity of depression, assessed by PHQ-9,^17^ and anxiety, assessed by GAD-7.^18^ Acceptability from the perspectives of male study participants and coaches who delivered the intervention was assessed in a nested qualitative study based on the seven component constructs of the theoretical framework of acceptability (TFA).^19^

### Sample Size

As BA is well-established as an effective intervention for less severe depression,^14^ ^20^ overall sample size was determined by pragmatic considerations with the aims of (1) benchmarking the pre-post results against the pre-post effects seen in the intervention and control arm of trials of BA delivered using a similar format; and (2) achieving meaning saturation (i.e. a comprehensive understanding of the issues raised in the data^21^) in the qualitative analysis of acceptability. We specified an enrolment target of 45 individuals as we believed this would achieve these aims.

### Data Collection

Study participants were invited to complete the PHQ-9 and GAD-7 questionnaires online via Qualtrics^22^ at baseline (pre-treatment), and 4- and 6-months. At baseline, participants were also asked to provide demographic information which included age, ethnicity, employer, occupation, and number of years worked in the NHS.

Qualitative data were collected in one-to-one semi-structured interviews conducted via telephone or video call (according to individual preference) with participants and coaches. Interviews explored experiences of the intervention and thoughts around content and delivery, including barriers and facilitators to following the treatment plan. Questions followed a topic guide developed by the study team informed by the TFA and input from the stakeholder advisory group, who provided guidance on the content and phrasing of questions. All interviews were recorded using an encrypted digital voice recorder and participants assigned an anonymous ID. Recordings were transcribed verbatim using a professional transcription service and uploaded to NVIVO-12.^23^

### Analysis

#### Quantitative analysis

Pre-post effect sizes and associated 95% confidence intervals (95% CI) were calculated for the PHQ-9 and GAD-7. A standardised mean difference, Hedge’s g is reported rather than a mean difference to facilitate comparison of the results with other studies. Separate effect sizes were calculated for baseline to four months and baseline to six months. For an effect size to be calculated, data was need for each time point on the measure. In the absence of a control condition, data were benchmarked against the CASPER trial.^24^ Although the CASPER trial has a different population (older adults) and setting (recruitment from primary care), it is comparable to the BALM study in a number of ways: it too uses a brief, guided self-help intervention, the intervention is based on Behavioural Activation principles, and the samples had subclinical levels of symptomatology.

#### Qualitative analysis

Qualitative data were analysed thematically using a Framework Approach.^25^ An a priori coding frame was developed based on acceptability/satisfaction with the intervention as defined by the TFA^19^ (figure 1), whilst also allowing for the emergence and exploration of any unanticipated themes. Initial analysis involved coding within each transcript followed by analysis across the codes and transcripts. Initial coding and analysis were conducted independently by K.B., H.S., P.G. and K.W., with team discussion to agree final analysis and identify discrepancies. Participant and coach data were analysed separately following the same analytical process.

**Figure 1.**
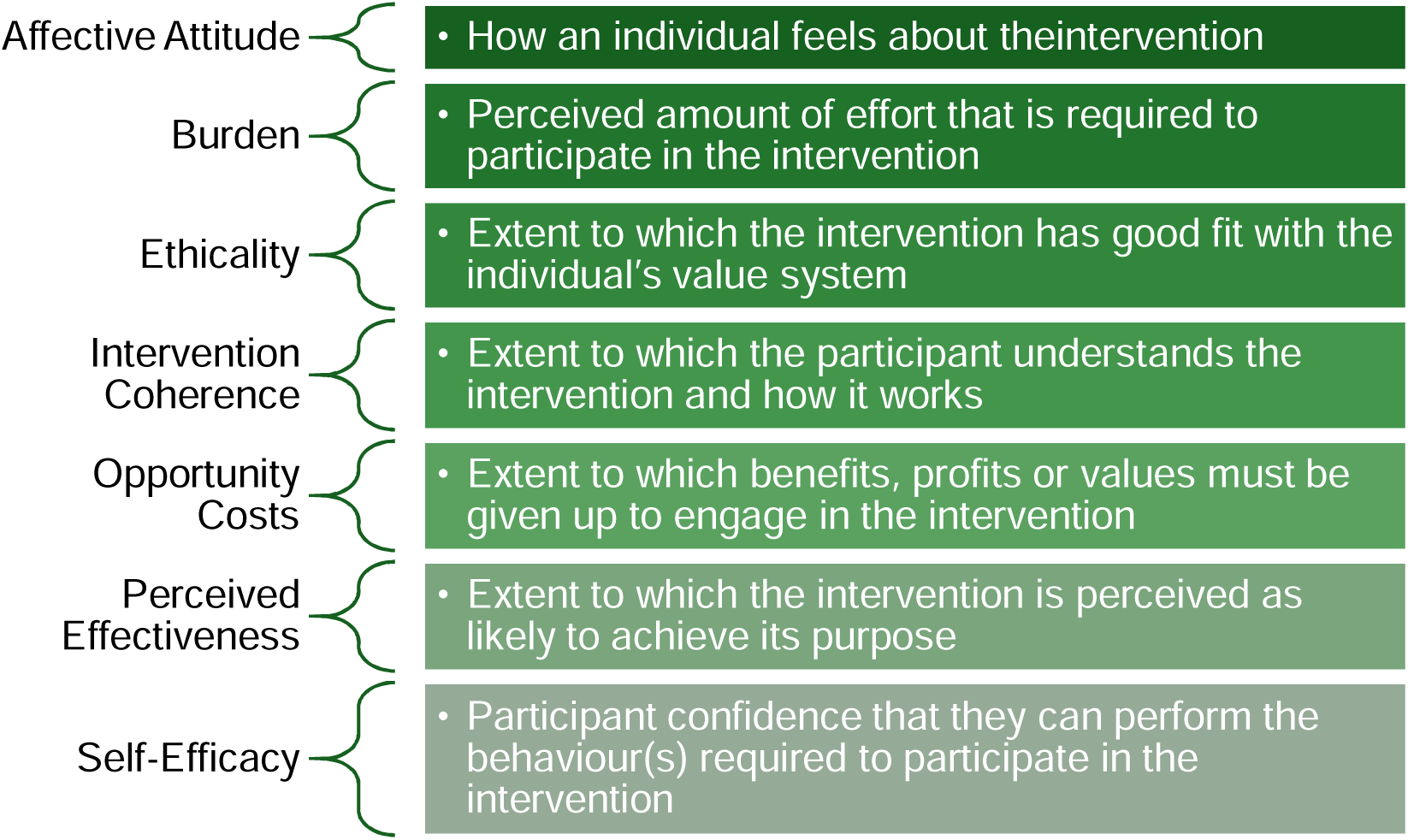
The seven component constructs of the TFA^19^.

## Results

### Recruitment and participant flow

Recruitment took place in the period January to September 2023. Two hundred and seventeen potential participants expressed interest in taking part in the study, of which 45 were enrolled (see figure 2). Recruitment ceased when the enrolment target of 45 individuals was reached. Of these, 30 completed the full BALM programme (‘completers’) and 15 did not complete the full intervention (at least 4 coach sessions; ‘non-completers’). This resulted in an intervention completion rate of 67% which is comparable to larger scale trials of BA in heterogeneous clinical populations.^14^

**Figure 2.**
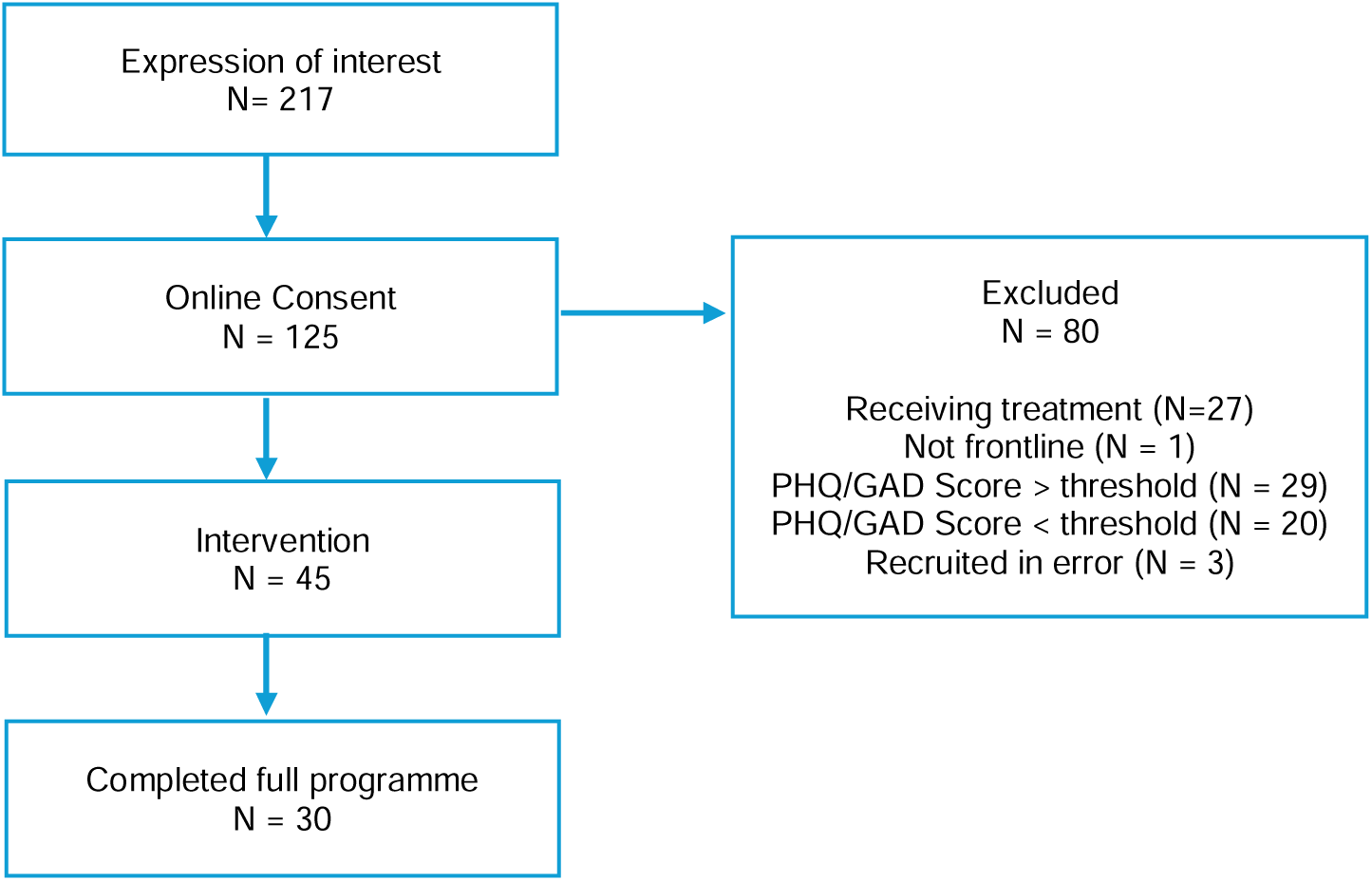
participant flow diagram.

#### Baseline data

Demographic and clinical characteristics (PHQ-9 and GAD-7) were gathered at baseline from male study participants. The mean age of the study participants was 40.8 years (SD = 11.0). All men self-identified as white/Caucasian. The majority of the men (61.9%) were married or in a domestic partnership. Male participants came from a range of NHS roles: nursing (N = 11), ambulance service team (N = 8), allied health professionals (N = 7), psychological professions (N = 6), management (N = 4), doctors (N = 3), health care support workers (N = 3) and other (N = 3). The mean number of years working in the healthcare sector was 10.5 (SD = 9.2).

The mean age of the coaches (N = 16) was 49.2 years (SD = 10.5). The majority were female (N = 10). All coaches self-identified as white/Caucasian. Coaches came from a range of NHS roles: allied health professionals (N = 6), admin and estates staff (N = 4), ambulance service team (N = 3), nursing (N = 1), management (N = 1), and healthcare science (N = 1).

#### Quantitative Outcomes

Table 1 summarises male study participant scores on the PHQ-9 and GAD-7 at baseline, 4 months and 6 months. Scores decreased from baseline to 4 months on both the PHQ-9 and GAD-7. While scores increased from 4 months to 6 months, the 6-month scores remained below those of the baseline scores.

**Table 1:**
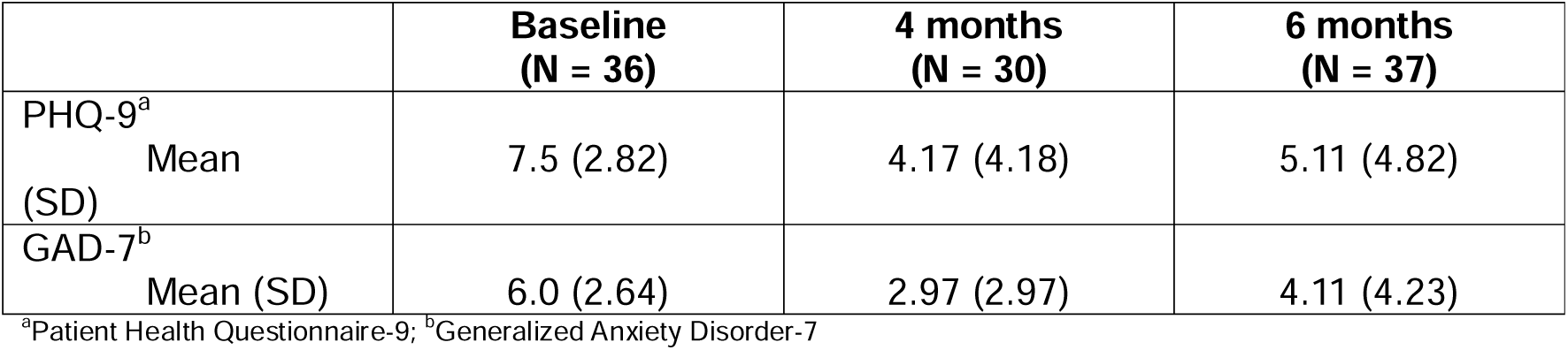
PHQ-9 and GAD-7 scores at baseline, 4 months and 6 months.

The pre-post effect size (Hedges’ g) at 4 months on the PHQ-9 was 0.86 (95% CI = 0.46 – 1.30) and at six months was 0.88 (95% CI = 0.51 – 1.28). For the GAD-7, the 4-month effect size was 1.0 (95% CI = 0.52 – 1.50) and at six months was 0.63 (95% CI 0.19 – 1.08). The 4-month figures were benchmarked against the 4-month pre-post effect sizes for the intervention and control conditions in the CASPER trial (see table 2).

**Table 2:**
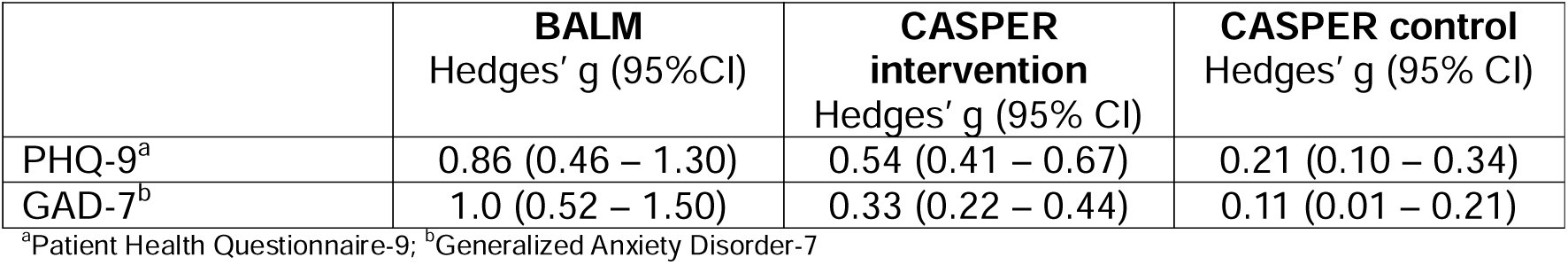
Benchmarking BALM pre-post effect sizes at 4 months with CASPER trial.

#### Qualitative Findings

In total, 32 study participants were selected via consecutive sampling and interviewed from the pool of 45 study participants who consented to take part in an interview at initial recruitment: 24 were ‘completers’ and 8 were ‘non-completers’. Twelve coaches were also selected for interview.

Acceptability of, and satisfaction with the intervention was investigated with the application of the TFA^19^ as a thematic analytical lens. The TFA defines acceptability as ‘a multi-faceted construct that reflects the extent to which people delivering or receiving a healthcare intervention consider it to be appropriate, based on anticipated or experienced cognitive and emotional responses to the intervention’. The framework consists of seven conceptual constructs related to the acceptability of interventions (figure 1). Findings from the TFA analysis incorporating both participant and coach perspectives are presented under each construct, below.

### Affective attitude

Affective attitude relates to how an individual feels about the intervention and was explored in terms of study participants’ and coaches’ views on receiving or delivering BALM, reasons for participating, and how individuals ‘felt’ about their overall experience. Evident in the data were a range of internal and external factors associated with participants’ and coaches’ attraction to the programme. For male study participants, prominent themes included recognising a need to do something about their mood, the appeal of a mental health programme with a specific focus on men, confidentiality, and the nature of the intervention on developing the ‘tools’ to help tackle work-related stress.

> *Because it said certainly with [the intervention] being around men, I’ve thought, oh, well, I’ll try and get involved in that, because I’ve really suffered in the past, there’s no two ways about that, I’ve been really down about life, about work…I thought, look, I could add another string to my bow, and how to cope and manage the stress that I get, that that would be great. (T0008)*

Several study participants described the appeal of the practical, action-oriented nature of BALM compared to more traditional talking-based therapies.

> *It wasn’t like counselling, it didn’t look like a sit and talk to a counsellor and someone will fix all your problems for you. It was more the fact that it was a tool that…even when the programme was finished, it was something that if it worked for me, I had nothing to lose, I might as well give it a go and see if it’s something that will help me and work. (Y0001)*

BALM coaches predominantly described positive affective attitude in relation to a desire to help others working in the NHS as well as the opportunity to develop their own knowledge, skills, and experience.

> *Ultimately what drives me is wanting to create a better quality of life for people, so addressing where there’s need and addressing where there’s areas where people’s quality of life isn’t so great. (CT24)*

### Intervention coherence

Intervention coherence was the extent to which study participants and coaches understood the BALM programme and how it worked. The overall aim of BALM and the principles of BA appeared to be well understood by participants. Key themes included the simplicity of the principles underpinning BA and the relationship between behaviour and emotional state, which resonated widely with men.

> *The booklet was fine, the booklet was good when I did read it…You know, I’d sometimes tell someone if I’d managed to do something this week, what I’ve tried to do for myself. [My coach] was quite clear that was the idea behind it, to support me. Looking at my decision to do things, was that impacting my mood, you know, for good or for worse. So, I’m totally on board with the idea behind it. (N0030)*
>
> *I found it really simple, really, really simple to follow. I think what I will take from the sessions that we did was that there was actually no need to go on and complicate matters…I think we just actually need to give the simple plans a good try, give it the opportunity to work. (T0028)*

All coaches commented positively on their training and how it had helped to develop a clear understanding of how Behavioural Activation works. Confidence and coherence of the intervention was enhanced by the provision of ongoing support from the research team and clinical supervision.

> *I felt confident, obviously, you feel a bit nervous, don’t you? But I felt confident that, do you know what, I can do this. I’ve got my resources. I’ve been on the training. I’ve got help. I’ve got the help [telephone] numbers if anybody is at risk on the appointment. (CY07)*

### Burden

The burden construct addressed the perceived amount of effort required to participate in BALM. The majority of male study participants who completed the full BALM programme did not perceive the telephone sessions with coaches to be a burden, with flexibility and the ability to schedule and/or reschedule sessions at convenient times a prominent theme in men’s experiences.

> *Really reasonable, actually, it [commitment to do intervention] wasn’t too much. And I think there was quite a lot of flexibility with the coach as well, so between us, we sort of discussed it. Do you want it to be weekly or the same day every week or…? So it was flexible, it was good…I can’t say there was [any burden], to be honest*. *(Y0001)*

However, the interviews with participants who did not complete the full programme reported that a lack of time and ability to commit to fitting in sessions with their coach over the duration of the programme was a common reason cited for withdrawing from the BALM sessions:

> *…so it’s just, it was really having to arrange the [coach] sessions and to do them and not having the time to do them that was the real issue… and I realised it just wasn’t going to work, so, you know, it wasn’t going to happen. (T0019)*

Undertaking ‘homework’ between telephone sessions, for example reintroducing meaningful activities such as physical exercise or seeing friends and family, was generally described by men as the main ‘effort’ required to participate in BALM, though this was not seen as burdensome.

> *I didn’t sacrifice, it was more of a positive intervention, a…positive things that you needed to make time to…to do things (T00043)*

Most coaches reported that they were unable to make adequate adjustments to their usual working role to accommodate their BALM coaching commitments. As such, finding sufficient time to prepare for sessions, maintain contemporaneous records, and schedule phone calls with participants was commonly highlighted by coaches to be burdensome, with many finding it impinged on their free time outside of work.

> *Sometimes I found it quite, just intrusive into my own free time. And sometimes it stopped me doing other things that I actually needed to do. I was having to, oh no, right, I’ve got to be around and I need to make sure I do this. And then when [a session] didn’t happen because I couldn’t get hold of people it’s oh God, right, well I’ll have to reorganise that for next week or I’ll have to try and reorganise that for tomorrow. (CY20)*

### Opportunity-costs

Opportunity costs are defined as the extent to which benefits, profits or values had to be given up to engage with the intervention. In terms of intervention delivery, there were mixed views in how study participants preferred to schedule phone calls with coaches to minimise opportunity costs. Some men accommodated calls during their working day, whilst others preferred to engage with BALM coaching sessions during their personal time. Several men described having no choice but to take calls outside of work because of the demands of their job role. As noted, flexibility in scheduling coaching sessions was key for minimising participant burden, particularly for participants who worked shifts. One participant discussed the difficulty in finding a mutually convenient time for sessions with his coach due to his inability to take calls at work, reflecting a similar theme prominent in analysis of data from coach interviews.

> *One of the things that I found really difficult… there was no way I could ever take phone calls [from my coach] during work because I couldn’t predict when I’d be needed to do things or, not need to do things. So, that then, it bled into my personal life, which is alright, but it was just really, really difficult to initially arrange appointments with my coach…It led to a point eventually where we couldn’t, our diaries didn’t match up anymore, so I had to get a new coach found. (T0017)*

Conducting sessions over the telephone was widely seen as beneficial by participants as it provided flexibility and convenience, but also confidentiality and a ‘safe space’ to talk that afforded a level of anonymity (see also Ethicality, below).

> *For me, [coaching sessions] over the telephone is a little bit easier…It’s a little bit, I don’t know how to describe it. A little bit kind of less personal, or you feel that you can, you know, be a little bit more maybe open or honest. With the phone as well, you don’t need to have internet connectivity. So, you’re not tied to just being in the house where you’ve got wi-fi or whatever it is…over the phone, you could be anywhere. You can go and drive off in your car and have 30 minutes…Because that’s what I did last week when I had it. I said to the Mrs, I said, right, I said, I’m going out. The kids are home. Are you all right looking after the kids just for, while I nip to B&Q? So, I went all the way to B&Q and just pulled over. [My coach] phoned us up, we had a chat, 30 odd minutes, and then I carried on to B&Q. (N0023)*

### Self-efficacy

Self-efficacy was defined as how confident male study participants and their coaches felt in undertaking the behaviours associated with the BALM process. Participants often discussed self-efficacy in relation to key BA principles such as increasing access to healthy behaviours; reducing avoidance behaviours; and understanding and attempting to address barriers to activation. Although not universal, awareness and understanding of the link between changing patterns of behaviour and improving mood was commonly highlighted in men’s accounts. For example, participants discussed being able to better recognise signs that they may be struggling with their mental health and behaviours that did not help their mood in the long term.

> *I’ve had a more open conversation with my wife around our sex life, I’ve reduced drinking, I’m very much more aware of some of lifestyle drinking choices which is not healthy and, yeah, the other stuff is basically the work things. (Y0009)*
>
> *One of the big things that I’ve noticed is just sometimes recognizing the signs that I am struggling. So, if I’ve had a particularly hard day at work, I would literally just slump in front of the TV and just try and switch off. And, from a mood point of view, that was probably the worst thing I could possibly do. So, now I can recognise that sign in myself and do something a bit more pro-actively. (T0017)*
>
> *I think I got to the stage where I was locking myself away. So often, when I was on night shifts and that as well, I’d sleep in a separate room from my wife…. And I find myself, if I’m waking up, even at a decent time, I’ll just be like, no, I’m staying in here. The kids are downstairs, the wife is downstairs and it’s a weekend and stuff, I don’t want to get involved in family activities because I’m just thinking, this is my little protective bubble. And I think that’s one of the things that I need to get out of, and that’s one of the things that I’m trying to do. I’m trying to kind of have more kind of family time really and trying to turn the family time into positive experiences rather than just the roller coaster of just levels of frustration. (N0023)*

Several coaches described how their confidence in delivering the intervention had increased as they became more experienced. Some also discussed adopting aspects of the intervention to benefit their own mental health. Although low self-efficacy was not a prominent feature of the data, there were some examples where coaches encountered difficulties in taking a collaborative approach to developing a treatment plan with participants.

> *Not that I was wanting to tell them what to do, but I was having to stop myself proposing different types of ideas, “have you thought about it…”, rather than getting them to do things. (CY20)*

### Ethicality

The ethicality construct related to what participants valued most or least about the BALM intervention and its ‘fit’ with their needs. For men who received the intervention, having the opportunity to address their mental health by focussing on behaviour change as opposed to ‘just talking’ was the most prominent feature of ethicality. For some men, the action-oriented nature of BALM also helped overcome the perceived stigma of engaging with mental health support. BALM being male-specific, confidential, and provided over the phone were also highly valued aspects of the intervention.

> *I think out of all the different therapies that are available men are most likely to engage with behavioural activation because it is something that they will not feel stigmatised by just discussing their behaviours and their activities. Whereas they may feel unable to open up. They may expect therapy to be sitting and crying and somebody holding your hand, sort of thing. I would suggest that of everything that I am aware of behavioural activation would be the most, well, maybe not the most helpful, but it would be the most likely to get people engaged in it. (T0028)*
>
> *I think the fact that it was over the phone, sort of, I don’t know, it helps in a way because it keeps that, sort of, distance. Well it did for me anyway, I just feel if I’d seen the, you know, seen the person then there’s the potential to then bump into him and just feel, or whatever. (Y0009)*

Frequently reported by coaches was a lack of mental health and wellbeing support for NHS staff in general, which BALM was perceived as helping to fill that gap. Negative views of the pressures of working in the NHS were often shared.

> *The NHS used to be a job to the grave. Now it’s putting people in graves. In my opinion. (CN01)*

### Perceived effectiveness

To determine perceived effectiveness, the extent to which BALM was perceived as likely to achieve its purpose, participants and coaches were asked about their views on the usefulness of the intervention, its potential viability in the wider NHS, and any recommendations for improvements. Both participants and coaches shared the view of the importance to provide BALM at scale in the NHS.

> *I think it should be rolled out more widely. I think it would make it more acceptable, and I think you get to help people a lot earlier, particularly from stuff that’s actually happened at work. ‘Cause of course, as I said, the time pressures at work, that if something does happen, and you feel mentally unwell because of it, the interaction to get something actually done, if you don’t actually go into your boss and say, right, I’m mentally unwell, then the debriefing and stuff like that, again, that’s such a challenge to try and get organised and to do. So I think if there was a bigger push with BALM to roll it out on a wider scale, it would be really useful. (Y0024)*
>
> *I’d be lying if I said it’s been a miracle cure…But it’s given me an outlet and it’s given me a mechanism of being able to speak to somebody who kind of gets me. Who is not, sorry, who is totally impartial. It’s not the wife who has got her own agenda with things and not the kids whose kind of wanting things…So, having somebody that’s impartial that I can speak to, that kind of gets it, has been really beneficial to me firstly. I think that’s helped me to change things, and recognise and think, you know. (N0030)*

Suggestions for improvement mostly centred on the development of technology associated with the intervention materials. Most relevant here were the diary and goal-setting activities in the BALM booklet/manual, with several participants suggesting accessibility could be improved with the development of an app to replace the need to write on paper. This was viewed by one participant as something that would also help with confidentiality.

> *I’m not great at picking up literature and, you know, using kind of booklets or keeping a diary and stuff. That might work for a lot of other different people. And so, if there’s any kind of different ways in which delivery of progress can be kind of monitored…Even if there’s like an app that could be developed where you can just, oh, today I’ll put in this activity. Rather than keeping a physical kind of paper file. Because, as much as I love my wife, I wouldn’t have wanted her nosing over my stuff, if you know what I mean. (N0023)*

## Discussion

In this study we evaluated the impact of a tailored, gender-responsive mental health intervention for improving depression and anxiety in male frontline NHS workers. We also sought participants’ and coaches’ views about the acceptability of the intervention. We found that the BALM intervention was acceptable for men with less severe depression and anxiety working in frontline NHS roles and contributed to reductions in depression and anxiety scores, which were sustained at 6 months follow-up. The benchmarking data indicated that the outcomes at 4 months compared well with another active intervention and clearly exceeded the level of improvement in the control condition of the randomised trial.

The study has several strengths. It is the first to evaluate the impact and acceptability of a BA intervention for improving depression and anxiety in men using a gendered lens, contributing novel findings of relevance wider than the study context. The use of the TFA to inform data collection and analysis enabled a theoretically informed, systematic assessment of the acceptability of the intervention. Rich data were gathered from qualitative interviews with a large sample of male participants and coaches who supported intervention delivery, including participant non-completers. The study also has important limitations. Participants were a self-selecting sample of men who may have been particularly motivated to take part in the intervention. It is possible their views on the acceptability of BALM are more positive than the wider population of male NHS staff. Our sample also lacked ethnocultural diversity and representation of some NHS staff groups, particularly ancillary roles, that limits the transferability of the findings. In accordance with a single sample pre–post design, we evaluated the effects of the intervention using standardised mental health measures. An inherent limitation of quasi-experimental research designs is they are not sufficiently reliable to make causal inferences. However, BA is a well-established intervention for depression and the pre-post results we achieved in the study are comparable against the pre-post effects seen in the intervention arm, and greater than those seen in the control arm, of high quality trials of BA delivered using a similar format.^20^ ^24^

Overall, our study findings highlight three key points of discussion. First, with levels of depression and anxiety rising in the NHS, and men’s lower participation in existing mental health programmes, our study shows the potential of BALM to help address the urgent need for early intervention and preventative mental health services outlined in the NHS Long Term Workforce plan.^26^ Most interventions implemented to date have had a focus on individual resilience with content including creative practices, emotion regulation activities, psychoeducation, and mindfulness.^3^ ^5^ However, these are likely to have limited efficacy as the overall resilience level of health care workers is already high.^27^ Our study has demonstrated that an evidence-based treatment programme for depression and anxiety can be tailored to be acceptable to, and beneficial for, male NHS frontline workers. Although interventions designed to alter individual health behaviours have received criticism as a reactive strategy to occupational stress that is more appropriately addressed through organisational change,^1^ ^3^ there is good evidence that common mental health problems are preventable and treatable in the workplace with appropriate individual intervention. Workers who receive treatment for stress, depression, and anxiety through workplace wellbeing interventions report higher levels of job satisfaction,^28^ and the NHS Long Term Workforce plan notes that investment of £80 per member of staff in mental health support can achieve net gains of £855 a year through savings from absenteeism and presenteeism.^26^

Second, our study is the first to show that tailoring a BA intervention using a gendered lens^16^ can yield benefit in men’s engagement with mental health support. BALM was perceived as acceptable by male participants across all constructs of the TFA. Tailoring interventions to make them more aligned with men’s needs and preferences has been widely suggested as a solution to men’s more reluctant attitude toward engaging with mental health services.^29^ Despite considerable research in this area, there remains little evidence on effective interventions that address these challenges. Our study has demonstrated the practical and action-oriented strategies of BALM were highly valued by and appealing to men, highlighting its promise as a mental health intervention for the broader male population. Our findings resonate with previous suggestions in the literature that collaborative approaches that are solution-focused, and allow men to enact an aptitude for problem-solving and self-management over passive dependency, can improve engagement.^11^ Other reported reasons for taking part in BALM were also consistent with factors reported in the wider literature on improving men’s engagement with psychological support. These included the appeal of a mental health programme focused on and marketed specifically for men, assurances on confidentiality and anonymity, and flexibility and convenience. Of particular relevance is the use of the telephone for intervention delivery. Coach sessions delivered over the telephone were considered convenient and highly acceptable by both male participants and coaches, replicating findings from previous studies of BA delivered in a similar format.^30^ In addition to offering further support to evidence that telephone-delivered mental health support does not negatively affect participant-coach interactions,^31^ our study suggests that it may actually enhance accessibility and the quality of therapeutic relationships for some male service users. However, our results do suggest that some minor modifications to our clinical protocol may further enhance acceptability. In particular, providing protected time to enable staff to accommodate their BALM coaching commitments, and the provision of tech-based (app) functionality for interactive elements of the intervention such as diary keeping, mood tracking, and activity scheduling.

Finally, on the basis of this initial study, BALM could have particular value as an occupational wellbeing provision outside the NHS, specifically in other frontline and emergency response communities such as police, military, and fire services, and in male-dominated occupations. Although there is high prevalence and diversity of men’s mental health challenges across a broad spectrum of workplaces and industries, the prevalence of depression within frontline and male-dominated workforces (>70% male employees) is substantially higher than in the general workforce.^9^ In addition to mental health risks arising from occupational characteristics of male-dominated and frontline roles, these work environments can foster “masculinity culture contests” that enhance social pressures to conform to gender norms that are associated with reticence to seek help, lower mental health literacy, and higher mental health-related stigma.^32^ ^33^ Evidence on effective early or preventative mental health interventions in these workplaces is limited, particularly those which incorporate gender transformative approaches that ‘‘work with and rework’’ traditional masculine norms.^16^ ^34^ The current study offers a platform for future research in these higher risk settings.

## Conclusion

This pre-post intervention study has shown that the delivery of a tailored, gender-responsive BA intervention was appealing to, and beneficial for, men working in frontline NHS roles with less severe depression and anxiety. The practical and action-oriented strategies of the intervention, and the confidential, flexible, convenient mode of delivery, worked to support men’s engagement with the intervention. The BALM intervention offers promise as a tailored workplace mental health programme that is aligned with men’s needs and preferences and can help overcome a reticence to engage with mental health support in NHS staff and beyond.

## Data Availability

All data produced in the present study are available upon reasonable request to the authors

## Footnotes

### Contributors

PG and DMc conceived, planned and designed this study and obtained the funding. Data collection was conducted by KB, HS, and KW. DMc conducted all quantitative data analyses. PG, KB, HS, and KW interpreted the qualitative data. PG drafted an initial version of the manuscript and drafted the final full manuscript. All authors contributed to the overall interpretation of results, ensuring the scientific rigour of the study, critically reviewed and contributed revisions to the manuscript, and approved the final version. The guarantor of the study is PG; accepts full responsibility for the finished work and/or the conduct of the study, had access to the data and controlled the decision to publish.

### Ethics approval

This study was given a favourable ethical opinion by Frenchay Research Ethics Committee on 28 Sept 2022 (REC reference: 22/SW/0113) and Health Research Authority approval on 29 Sept 2022 (IRAS project ID: 314095). Participants gave informed consent to participate in the study before taking part.

### Patient and public involvement

Patients and/or the public were involved in the design, conduct, reporting or dissemination plans of this research. Refer to the Methods section for further details.

